# Perinatal Outcomes of Fetuses and Infants Diagnosed with Trisomy 13 or Trisomy 18

**DOI:** 10.1101/2021.10.08.21264249

**Authors:** DonnaMaria E. Cortezzo, Leandra K. Tolusso, Daniel T. Swarr

## Abstract

**Objectives:** To identify factors associated with adverse prenatal, perinatal and postnatal outcomes, and determine the utilization medical care for fetuses & infants with trisomy 13 (T13) and trisomy (T18).

**Study Design:** This population-based retrospective cohort study included all prenatal and postnatal diagnoses of T13 or T18 in the Greater Cincinnati area from 1/1/12-12/31/18. Overall survival, survival to hospital discharge, approach to medical management, and maternal, fetal and neonatal characteristics are analyzed.

**Results:** 124 pregnancies (125 fetuses) were identified, which resulted in 72 liveborn infants. Male fetal sex and hydrops were associated with a higher rate of spontaneous loss. The median length of survival was 7 days (95% CI, 0-18 days) and 29 days (95% CI, 2-115 days), for infants with T13 and T18, respectively. Of the 27 infants who were alive at 1 month of age, 14 (52%) were alive at 1 year of age. Only the trisomy type, chosen goals of care (comfort care), and extremely low birthweight were associated with overall length of survival. A high degree of variability existed in the use of medical services, with 28% of infants undergoing at least one surgical procedure and some children requiring repeated (up to 29) or prolonged hospitalization (> 1 year).

**Conclusions:** Although many infants with T13 or T18 did not survive past the first week of life, up to 25% of infants lived for more than one year. Length of survival for an individual infant cannot be easily predicted, and surviving infants have high health care utilization throughout their lifespan.

## Introduction

Trisomy 13 (T13) and Trisomy 18 (T18) are among the most common fetal life-limiting diagnoses, with a combined prevalence of roughly 1 in 1800 pregnancies, or 1.68 per 10,000 births for T13 and 4.08 per 10,000 births for T18.^1-4^ Each condition has previously been referred to as a lethal diagnosis, with near universal descriptions of profound neurodevelopmental impairment. Historically, invasive medical interventions were considered by many to be “futile” and a “comfort care” approach was the only option presented to families. Recent research studies and debate within the medical literature have challenged these views, along with the increasingly recognized diverse perspectives of families with children affected by trisomy 13 and trisomy 18.^5-9^ A growing number of medical institutions are offering a range of medical and surgical interventions for children with trisomy 13 or 18, and families are increasingly electing to pursue these interventions for their children.^10, 11^

There is significant variability in the choices families make regarding prenatal and postnatal care and the medical care delivered by providers following a diagnosis of T13 or T18.^12-14^ Some families choose to continue the pregnancy, with most hoping to meet their child alive. After birth, some choose to focus on the comfort of their child and the time they have available to spend together as a family. Other elect intensive medical care with the goal of prolonging the duration of life, even if this may entail painful medical procedures or impact the quality of the time the parents are able to spend with their infant. Irrespective of the goals of care, families appreciate balanced and personalized information, and want healthcare providers that respect their choices and provide support throughout this process.^7, 15-17^

To meet these needs for families, it is important providers understand outcomes of pregnancies and neonates with T13 or T18. Rates of spontaneous pregnancy loss are variable, with reports ranging from 32%-87%. Intrapartum deaths and fetal intolerance of labor are high, with rates of cesarean section reported from 64%-80%.^18^ Although the median survival of infants with T13 or T18 is one to three weeks, some children with these diagnoses live for years.^19^ Previous population-based studies have reported median survival of 7-10 days for neonates with T13 and 10-14.5 days for neonates with T18, but 46% of birth admissions are discharged alive and survival to 5 years occurs in 9.7% and 12.3% of infants with T13 and T18, respectively.^19-23^ Therefore, the universal application of the term “lethal” for these diagnoses is not appropriate, and an increasing number of individuals with T13 or T18 are receiving medical interventions, including surgical procedures.^23, 24^

The variation in pregnancy outcomes, survival outcomes, treatment, and goals of care is likely due in part to variation in counseling, care practices, and medical interventions offered to individuals with T13 or T18. These factors are all interrelated, with the evolution in approaches to medical management that are offered to families and the variability in outcomes making it challenging for healthcare providers to counsel families with the most accurate and up-to-date information.^9, 17, 20, 25-28^ The objective of this retrospective cohort study of pregnancies and neonates with a diagnosis of T13 or T18 in the greater Cincinnati region over a 7 year period was to identify factors associated with adverse prenatal, perinatal and postnatal outcomes, and determine the utilization medical care for fetuses and infants with T13 and T18 in order to better understand these unique patients and facilitate high quality counseling and shared decision-making.

## Methods

### Design

This study was approved by the Cincinnati Children’s Hospital Institutional Review Board (IRB), as well as each IRB for all hospitals with obstetrics services across the Greater Cincinnati Region. Subjects were identified through genetic counselors, regional cytogenetics laboratories, referrals to perinatal hospice, and EPIC data query at each institution providing obstetric and newborn services for neonates with an ICD code of T13 or T18 and pregnancies complicated by a diagnosis of T13 or T18 between January 1, 2012 and December 31, 2018. All prenatal and postnatal diagnoses of T13 or T18 at all delivery hospitals in the Greater Cincinnati metropolitan region [1 level IV Neonatal Intensive Care Unit (NICU), 4 level III NICUs, and 10 level I/II delivery hospitals] were included in an attempt to minimize ascertainment bias. We selected the 2012-2018 epoch so that the treatment decisions offered to and chosen by families would reflect current clinical practice. The diagnosis was confirmed by manual review of the cytogenetics report, and only subjects with “full” trisomy 13 or 18 were included. Subjects were excluded from the study if: 1) Medical records were not available for review; 2) All of the care after diagnosis was provided outside of the region; or, 3) Chromosomal mosaicism or more complex chromosomal rearrangements were present. A detailed retrospective chart review was performed to determine outcomes of pregnancies and neonates with a prenatal or postnatal diagnosis of T13 or T18.

### Data Collection

Manual data extraction was performed from a complete review of all charts meeting inclusion criteria. Data was directly entered into a REDCap electronic database, and consisted of 153 distinct variables spanning demographic, maternal, prenatal, perinatal, and postnatal outcomes. The National Death Index was also accessed to verify the live-born patients who had died and ascertain the reported cause of death.

### Data Analysis

De-identified data was exported from REDCap and all analyses were performed using the R statistical software package. Descriptive statistics were calculated using the “Rcmdr” package. Survival was calculated from birth and death dates. Living children were censored on the day that their last chart review and National Death Index search was performed. Univariate and multivariate Cox regression analysis of factors associated with duration of survival was performed using the “survival” R package; reported p-values reflect the Wald test. Kaplan-Meier survival plots were also generated with the “survival” package. Associations between two categorical variables were tested using Fisher’s exact test, and associations between two groups with continuous variables were tested using the Wilcoxan rank-sum test. Differences between groups were considered statistically significant when the *p-*value was less than 0.05. The World Health Organization criteria for classification of birthweights and prematurity were used. Where appropriate, the Centers for Disease Control definition was applied to classify structural abnormalities as a “major congenital malformation” when the “structural changes⃛have significant medical, social or cosmetic consequences for the affected individual, and typically require medical intervention.”^29^ Structural anomalies were considered a “major surgical malformation” when the standard of care (for otherwise healthy infants) typically involves surgical repair in early infancy (e.g. congenital diaphragmatic hernia, gastroschisis, omphalocele, myelomeningocele).

## Results

A total of 125 fetuses or infants diagnosed with T13 (n = 38) or T18 (n = 87) were identified. (Table 1) The total birth prevalence (livebirths, spontaneous abortions <20 weeks, intrauterine fetal demises at and above 20 weeks, and elective termination of pregnancy) for T13 was 1.62 per 10,000 livebirths and the prevalence among livebirths was 1.00 per 10,000 livebirths. The total birth prevalence for T18 was 3.76 per 10,000 livebirths and the prevalence among livebirths was 1.86 per 10,000 livebirths. The mean maternal age was 33 years (range 14-46), and the mean gravida was 3 (range 1-14). Most mothers identified as Caucasian (79%), with complete demographic data outlined in Table 1. Just under half of all mothers had at least one prior pregnancy loss, and 5% reported a previous pregnancy affected by chromosomal abnormalities.

**Table 1.**
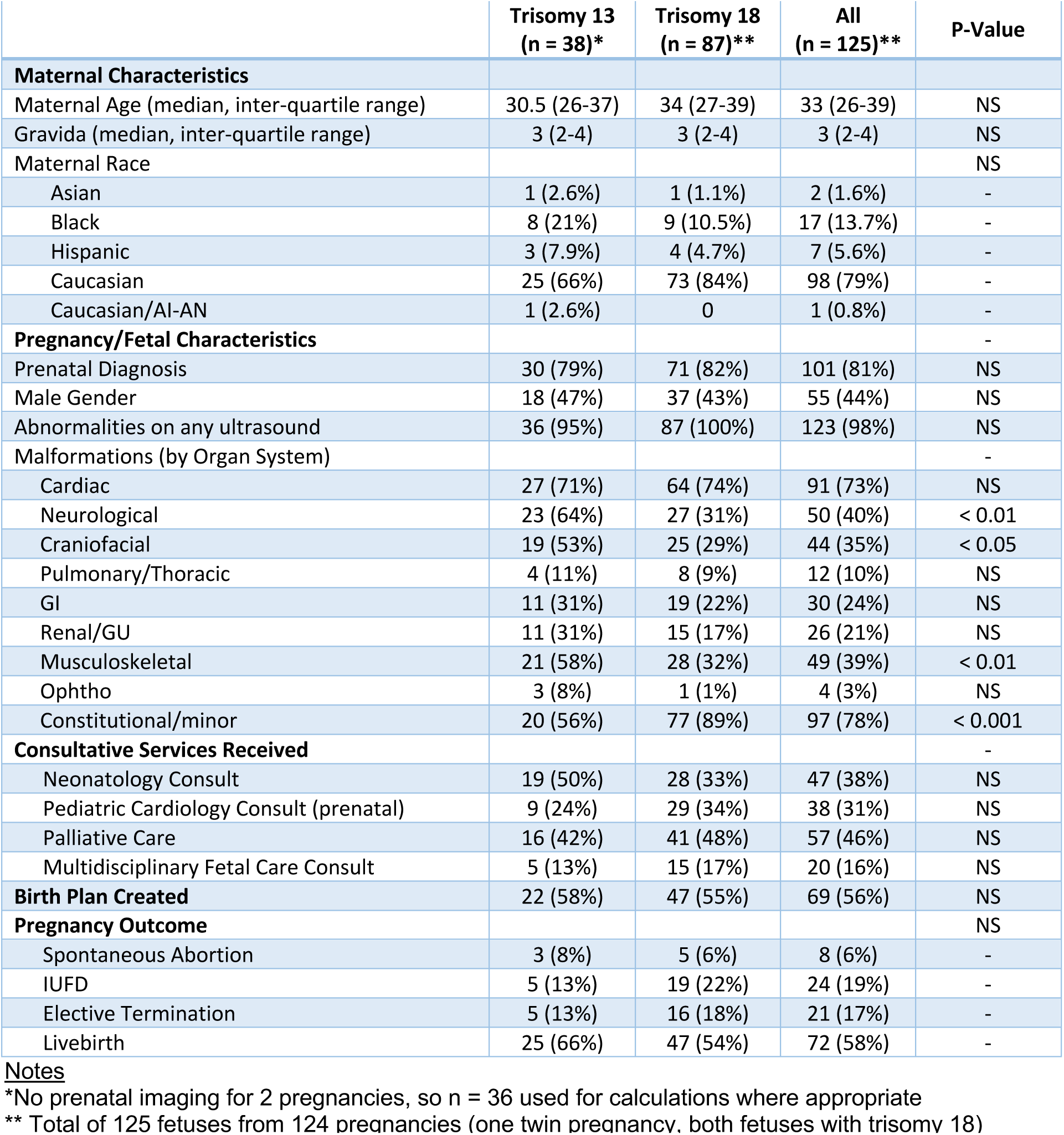
Prenatal Characteristics.

The majority of diagnoses were made during the prenatal period (101/125, 81% overall) - 30/38 (79%) for T13, 71/87 (82%) for T18. In total, 88 women (71%) underwent amniocentesis (1 twin pregnancy, both twins affected with trisomy 18), 11 (9%) underwent chorionic villus sampling, and 1 had a karyotype performed after a spontaneous abortion. Of the 24 infants diagnosed after birth, two had a false-negative cell-free fetal DNA screen (both T18), one had a maternal serum quad screen that was reported as high-risk for trisomy 21 (with no follow-up invasive testing), one had a normal prenatal ultrasound without any additional prenatal testing, and in the remaining 8 pregnancies families either declined prenatal diagnostic testing despite high-risk genetic screening results and/or ultrasound findings, or had noninvasive testing that technically failed. At least one abnormality was seen on prenatal imaging in 98% of pregnancies. The number of organ systems with at least one structural malformation seen on prenatal ultrasonography was higher for fetuses with trisomy 13. (Table 1) The incidence of congenital heart disease was similar between the two groups (71% versus 74% for trisomy 13 versus 18, respectively). The specific congenital heart lesions identified during the prenatal period are listed in Supplemental Table 1.

Of the 125 pregnancies included in this cohort, 6% ended following a spontaneous loss before 20 weeks gestation (n = 8), 19% ended with a loss during or after the 20^th^ week gestation (intrauterine fetal demise; n = 24), 17% chose to terminate the pregnancy (n = 21), and 58% resulted in a liveborn infant (n=72). In univariate analyses, male fetal sex, lower maternal gravida, and the presence of hydrops were associated with higher incidence of spontaneous pregnancy loss. The percentage of fetuses with prenatally diagnosed congenital heart disease was higher among the pregnancies resulting in a liveborn infant than those that resulted in a spontaneous loss. (Supplemental Table 2) We sought to determine whether the counseling documented during the prenatal period was associated with the ultimate pregnancy outcome, and the overall length of survival or survival to initial hospital discharge after birth (for liveborn infants). Less than half of parents met with either a neonatologist (38%), pediatric cardiologist (31%), multidisciplinary fetal care team (16%) and/or pediatric palliative care specialist during the prenatal period. (Table 1) An additional 13% (n=16) met with Palliative Care after delivery. Compared to women whose pregnancy ended in a livebirth or spontaneous loss after 20 weeks gestation, women who chose to terminate their pregnancy were significantly less likely to have had a prenatal consultation with a Neonatologist, Pediatric Cardiologist, or Pediatric Palliative Care Specialist. (Supplemental Table 3) A slight majority (56%; n = 69) of families had a birth plan in place at the time of delivery.

Birth and delivery data are summarized in Table 2, and postnatal care outcomes are depicted in Supplemental Figures 1-2. Of the 125 fetuses included in the cohort, 72 (58%) were born alive. Overall, nearly half of infants were born by Cesarean section, though this rate was higher for infants with trisomy 18 (60%) compared to infants with trisomy 13 (28%). Half of the women in the cohort (n = 36) either had scheduled deliveries or went into labor spontaneously. Ten women (14%) went into preterm labor, and 11 women (15%) had premature rupture of membranes. 17% of women (n = 12) were delivered for maternal indications (e.g. preclampsia), and an additional 17% were delivered due to concern for fetal well-being. Infants with trisomy 18 weighed significantly less than infants with trisomy 13 at birth (median birthweight 1851g versus 2286g), even though the median gestational age of the two groups was not significantly different. Regardless of the location of birth, just under half (44%) of all infants were ultimately admitted to a level III NICU, but 92% of infant-family dyads were offered a full resuscitation at the time of delivery. In the six cases (8%) where invasive interventions were not offered, major anomalies or other comorbidities (e.g. proboscis, hydrops, extreme prematurity) were present that were felt to be incompatible with survival. Nearly half (47%) of infants received positive-pressure ventilation (PPV) in the delivery room and 8% received endotracheal tube placement. No infants received chest compressions, emergent IV access, or cardiac medications. Families were nearly divided in either birthweight less than 1000g (extremely low electing a comfort care approach after birth (47% overall) or choosing to pursue invasive treatment with the goal of extending life (43% overall). The remaining 10% of families chose a trial of non-invasive therapies, such as nasal cannula and/or nasogastric (NG) feeds. (Table 2)

**Table 2.** Characteristics & Delivery Room Management of Liveborn Infants.

|  | **Trisomy 13**<br>(n = 25) | **Trisomy 18**<br>(n = 47) | **All**<br>(n = 72) |
| --- | --- | --- | --- |
| **Birthweight** (median, IQR) | 2245<br>(1900-2857) | 1890<br>(1572-2115) | 1976<br>(1605-2295) |
| **Gestational age** (median, IQR) | 36 6/7<br>(34 2/7 - 38 2/7) | 37 2/7<br>(36 0/7 - 38 2/7) | 37 1/7<br>(35 3/7 - 38 2/7) |
| **Mode of Birth** |  |  |  |
| Vaginal delivery | 18 (72%) | 19 (40%) | 37 (51%) |
| Cesarean section | 7 (28%) | 28 (60%) | 35 (49%) |
| **Location of Birth** |  |  |  |
| Level I/II Facility | 4 (16%) | 16 (34%) | 20 (28%) |
| Level III Facility | 21 (84%) | 31 (65%) | 52 (72%) |
| **NICU Admission** | 9 (36%) | 23 (49%) | 32 (44%) |
| **Offered Attempt at DR Resuscitation** | 23 (92%) | 43 (91%) | 66 (92%) |
| **Initial Treatment Decision** |  |  |  |
| Comfort care | 12 (48%) | 22 (47%) | 34 (47%) |
| Trial of non-invasive therapies | 11 (44%) | 20 (43%) | 31 (43%) |
| Full intensive care | 2 (8%) | 5 (11%) | 7 (10%) |
| **Delivery Room Interventions** |  |  |  |
| Blow-by O2 | 3 (12%) | 9 (19%) | 12 (17%) |
| Positive-pressure ventilation (PPV) | 6 (24%) | 28 (60%) | 34 (47%) |
| Intubation | 6 (16%) | 2 (4%) | 6 (8%) |
| **Select Congenital Malformations** |  |  |  |
| Congenital Diaphragmatic Hernia (CDH) | 1 (4%) | 3 (6%) | 4 (6%) |
| Myelomeningocele (MMC) | 2 (8%) | 2 (4%) | 4 (6%) |
| Anorectal malformation | 1 (4%) | 2 (4%) | 3 (4%) |
| Omphalocele | 2 (8%) | 3 (6%) | 5 (7%) |
| Gastroschisis | 0 | 0 | 0 |
| Cleft lip/palate | 9 (36%) | 7 (15%) | 16 (22%) |
| Holoprosencephaly | 8 (32%) | 3 (6%) | 11 (15%) |

**Notes**
DR – Delivery Room
IQR – Inter-quartile range

The median length of survival was 7 days (95% CI, 0-18 days) and 29 days (95% CI, 2-115 days), for infants with trisomy 13 and trisomy 18, respectively. (Figure 1A-B) Of the 27 infants with either trisomy 13 or 18 who were alive at 1 month of age, 14 (52%) were alive at 1 year of age, with a median survival for this group of 590 days. In univariate analyses, the type of trisomy (trisomy 13), birthweight, ELBW), very or extremely low-gestational age (VLGAN/ELGAN), and choosing a comfort-measures only approach, were associated with a shorter length of survival after birth. (Figure 1C-F, Supplemental Table 4) In multivariate regression analysis, only the goals of care, trisomy type, and birth weight of < 1000g remained significantly associated with the length of survival. (Table 3) Notably, neither the number of organ systems impacted by one or more malformations nor the presence of congenital heart disease were associated with length of survival. The total number of individual major congenital malformations was too small to permit a detailed statistical analysis of outcomes by each type of malformation. However, malformations that were never associated with survival beyond the first week of life or survival to hospital discharge in this dataset included congenital diaphragmatic hernia (n = 4, death on DOL #0 in all cases) and hypoplastic left heart syndrome (n = 5, death on DOL#0-7). Moreover, all infants born at less than 32 weeks gestation or insufficiency and apnea were frequent less than 1000g died within the first week of life and prior to hospital discharge. There was no significant association between myelomeningocele or cleft lip/palate on length of survival. (Supplemental Figure 3) Prenatal consultation with Neonatology, Fetal Care, or Palliative Care was not associated with a change in the overall duration of survival for liveborn infants. However, consultation with a pediatric cardiologist during the prenatal period was associated with an increased length of survival. (Supplemental Table 2)

**Figure 1.**
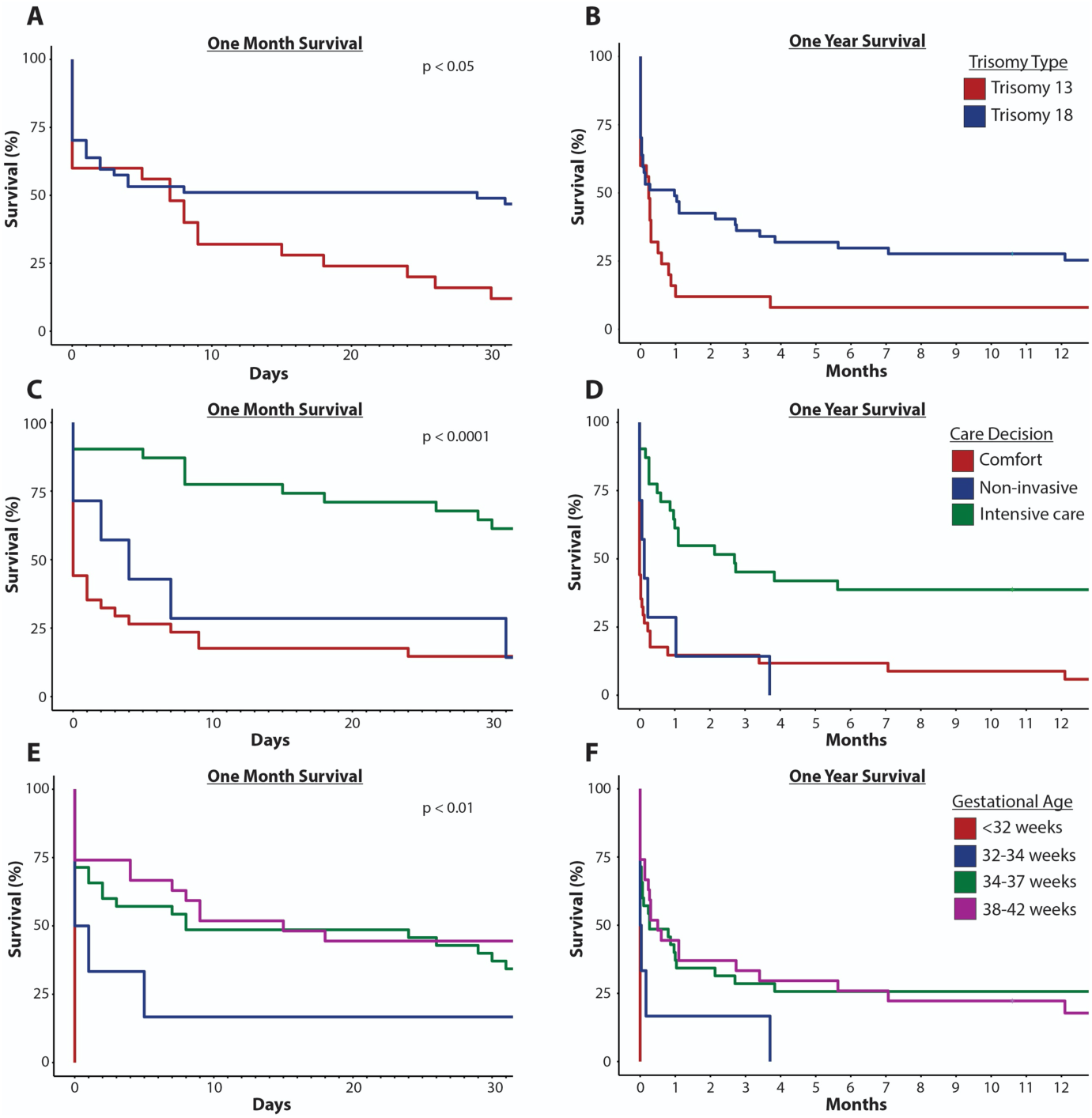
Survival for Liveborn Infants with T13 or T18. (**A-B**) Kaplan-Meier curves for infants with T13 (red) or T18 (blue) are shown for 30 day (A) and 12-month (B) intervals. (**C-D**) Kaplan-Meier curves stratified base on the care decision made (comfort care, red; non-invasive support, blue; intensive care, green) for 30 day (C) and 12-month (D) intervals. (**E-F**) Kaplan-Meier curves stratified base on gestational age at birth for 30 day (C) and 12-month (D) intervals.

**Table 3.**
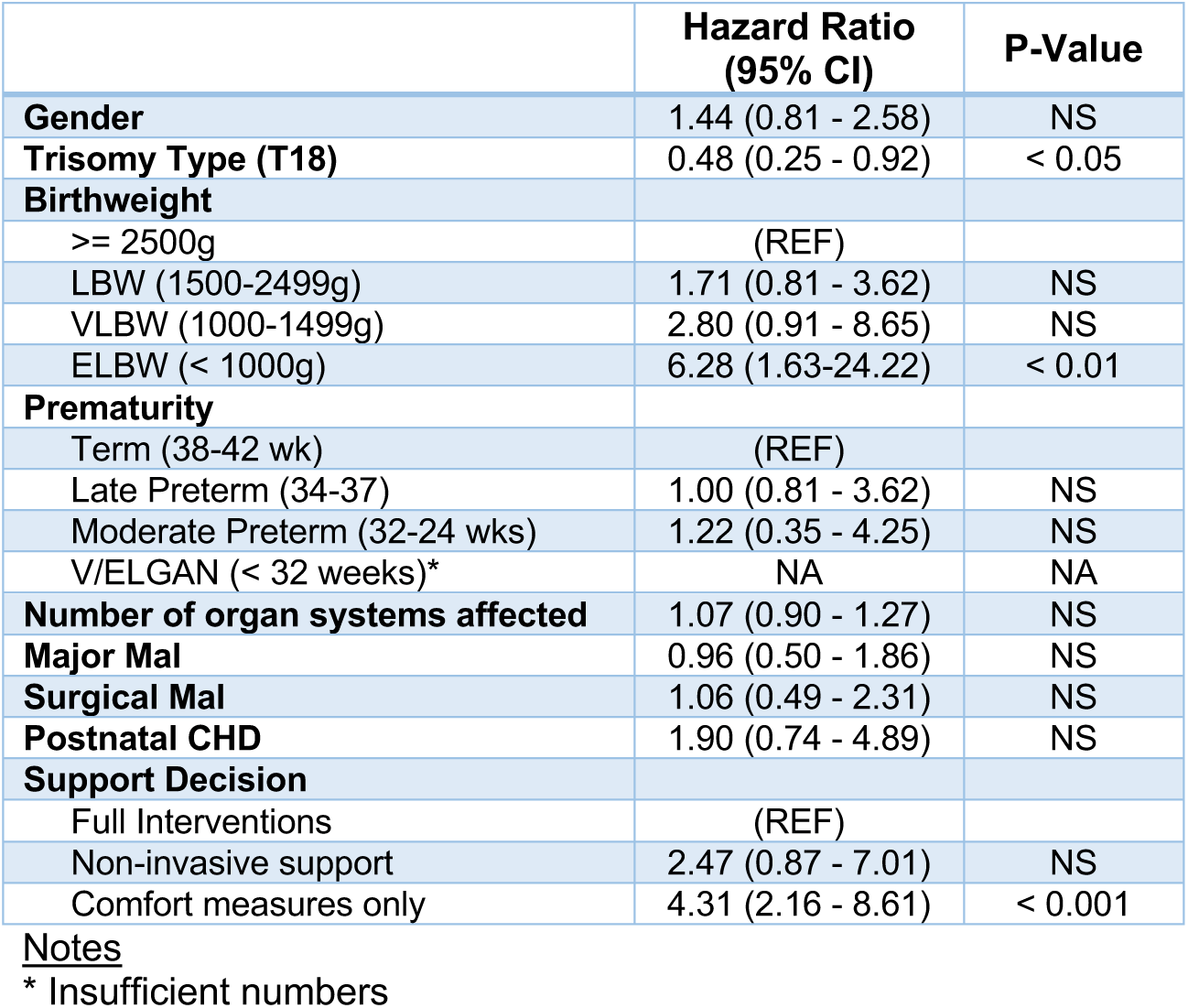
Factors Associated with Survival.

|  | Hazard Ratio<br>(95% CI) | P-Value |
| --- | --- | --- |
| **Gender** | 1.44 (0.81 - 2.58) | NS |
| **Trisomy Type (T18)** | 0.48 (0.25 - 0.92) | < 0.05 |
| **Birthweight** |  |  |
| >= 2500g | (REF) |  |
| LBW (1500-2499g) | 1.71 (0.81 - 3.62) | NS |
| VLBW (1000-1499g) | 2.80 (0.91 - 8.65) | NS |
| ELBW (< 1000g) | 6.28 (1.63-24.22) | < 0.01 |
| **Prematurity** |  |  |
| Term (38-42 wk) | (REF) |  |
| Late Preterm (34-37) | 1.00 (0.81 - 3.62) | NS |
| Moderate Preterm (32-34 wks) | 1.22 (0.35 - 4.25) | NS |
| V/ELGAN (< 32 weeks)* | NA | NA |
| **Number of organ systems affected** | 1.07 (0.90 - 1.27) | NS |
| **Major Mal** | 0.96 (0.50 - 1.86) | NS |
| **Surgical Mal** | 1.06 (0.49 - 2.31) | NS |
| **Postnatal CHD** | 1.90 (0.74 - 4.89) | NS |
| **Support Decision** |  |  |
| Full Interventions | (REF) |  |
| Non-invasive support | 2.47 (0.87 - 7.01) | NS |
| Comfort measures only | 4.31 (2.16 - 8.61) | < 0.001 |

Notes
\* Insufficient numbers

Just under half of all liveborn infants were eventually discharge home (n = 32, 44%). When limited to the 32 infants admitted to a NICU, two-thirds were eventually discharged home. The number of infants with trisomy 18 discharged home from the NICU (74%) was higher than for infants with trisomy 13 (44%). (Supplemental Table 6) Overall, length of stay in the hospital after birth was a median of 16 days (range, 1-531 days). Of the 32 patients discharged home from the hospital after birth, nearly half were readmitted to the hospital at least once (range: 1-29 re-hospitalizations). The total number of days spent in the hospital during these readmissions varied widely, ranging from 1-713 days. Respiratory morbidities contributing to both the need for and overall length of hospitalization. Most infants in the cohort were diagnosed with either central, obstructive, or mixed apnea at some point in their life (88%). Just under half of children required some degree of respiratory support (47%), and a quarter (24%) required mechanical ventilation. (Supplemental Table 6) The median time spent on a mechanical ventilator was 8 days (range 1-515 days). A tracheostomy was placed to facilitate long-term ventilatory support in ten children (14%). All neonates who received invasive interventions required a nasogastric tube or gastrostomy tube for at least some of their enteral nutrition. More than a quarter (28%) of children with trisomy 13 or 18 underwent at least one non-cardiac surgical procedure at some point in their life (median 3, range 1-17), at a median age of 233 days (range 0-2,173 days). The most common procedures performed were gastrostomy tube and tracheostomy placement. (Supplemental Table 3) Just under half of liveborn infants (n = 35, 49%) had a postnatal echocardiogram. Significant cardiovascular pathology was identified on all but one study. However, congenital heart disease was suspected during the prenatal period in 90% of liveborn infants, based on the level II ultrasound and/or fetal echocardiography. The most common findings were ventricular septal defects, atrial septal defects, polyvalvular abnormalities, patent ductus arteriosus, double outlet right ventricle, and tetralogy of Fallot. (Supplemental Table 8) Most liveborn infants with congenital heart disease confirmed by echocardiography (60% for T13, 72% for T18) developed cardiac symptoms at some point in their life and required management with medications (e.g. diuretics). Eighteen of these infants (51%; T13 50%, n=5; T18 52%, n=13) had significant pulmonary hypertension. No children with trisomy 13 received invasive cardiac interventions in our cohort. Cardiac surgery was considered in 6 children with trisomy 18. One child was ultimately determined to not be a candidate for surgical repair after cardiac catheterization revealed pulmonary hypertension. The other five children underwent surgical repair for a patent ductus arteriosus, ventricular septal defect, atrial septal defect, and/or patent foramen ovale. No children underwent surgical intervention for more complex congenital heart lesions.

## Discussion

This large regional retrospective cohort provides a detailed overview of the prenatal, perinatal and postnatal outcomes of fetuses and infants with trisomy 13 and 18. To our knowledge, such a comprehensive and inclusive study of both prenatal and postnatal outcomes has not previously been reported in the literature.

There is significant variation in the reported outcomes of pregnancies involving a fetus with trisomy 13 or 18, likely in part reflecting the changes in choices made by families and medical providers over time. Anywhere from 2%-55% of families elect for pregnancy termination.^14, 18, 30^ For those who elect to continue their pregnancies, the rates of spontaneous abortions or fetal demise remain high.^3, 18, 31, 32^ However, our data highlight that although there is an increased risk of spontaneous pregnancy loss in these pregnancies, the majority of pregnancies resulted in the delivery of a liveborn infant. Male sex and the presence of fetal hydrops were the only factors associated with a higher incidence of spontaneous loss. These data highlight a key point for counseling that regardless of the family’s goals for postnatal care, many of these families will meet their infant alive, and both obstetric and pediatric care providers need to be prepared to guide families through this process.

Naturally, families cannot make care decisions or adequately prepare for the neonatal period of their child’s life without an accurate prenatal diagnosis. In our cohort, a significant concern was raised for aneuploidy in all cases where both non-invasive maternal blood screening (either cell-free DNA or serum screening) and level II ultrasonography was performed. However, in a small number of cases, cell-free DNA screening reported a low-risk result or no abnormalities were seen on ultrasound. As such, it remains critical that families understand the limitations of these screening tests during the prenatal period. Although many families in our cohort elected to obtain diagnostic testing with an amniocentesis or chorionic villus sampling, some families deferred diagnostic genetic testing until after birth. Even in the absence of a definitive diagnosis, simply establishing a high suspicion for either trisomy 13 or 18 may impact goals of care for the pregnancy. In one study, patients who elected for a comfort measure only approach to care after prenatal diagnosis of T13 or T18 were more likely to deliver vaginally and have an elective induction of labor than those who elected for neonatal intervention.^33^ Other studies have reported a much higher rate of caesarian delivery.^18^ In our cohort, the rate of Cesarean delivery for infants with trisomy 13 was comparable to the national average (28% versus 31.7%), but was much higher for infants with trisomy 18 at nearly 60% of deliveries.^34^

A number of recently published large studies have provided significant insights into survival rates of infants with trisomy 13 and 18, but interpreting these data in the context of the highly varied treatment pathways that families choose for their infants remains complicated.^5, 6, 35, 36^ In our cohort, the median survival time of 7 days (95% CI, 0-18 days) for infants with trisomy 13 and 29 days (95% CI, 2-115 days) for infants with trisomy 18 are consistent with recently published survival statistics.^19^ However, it is clear from both our data and the recently published literature that a significant minority of infants with both trisomy 13 and trisomy 18 live well-beyond the neonatal period, with some children living for many years.^19 20, 21^ Also consistent with previously published data, we found that prematurity and low birth weight were associated with a decreased length of survival, particularly for very or extremely premature infants (< 32 weeks gestation) and/or extremely low birthweight (< 1000g). We speculate that the lack of significant effect on mortality for more modest decreases in birthweight may be related to the relatively high incidence of IUGR/SGA, particularly among infants with trisomy 18, which may not directly impact mortality. In our study, infants with trisomy 18 had overall higher rates of survival compared to infants with trisomy 13. This is consistent with some, but not all other reports in the literature.^6, 37, 38^

The factor most strongly associated with overall length of survival in our study was the level of support chosen by families and their care providers. Compared to infants who received all intensive care as medically indicated, infants who received non-invasive support or comfort care only were more than twice as likely to die (hazard ratios of 2.47 and 4.31, respectively). Even though these analyses corrected for gender, trisomy type, birthweight, gestational age, and the presence of major congenital malformations, major surgical malformations, and congenital heart disease, it is highly likely that these observed associations are still highly confounded by additional medical morbidities and medical factors that impact families’ and medical providers’ care decisions. For example, no infants in our cohort with either CDH or HLHS survived past the first week of life or to hospital discharge – the very reasonable limitations of care made in these cases should not be interpreted to mean that had interventions been offered, the length of survival would have been improved. It is also worth noting that these results are in contrast to a previous study that reported while parental goals of care altered the treatment intensity for patients, it did not affect the length of survival. ^39, 40^ It is possible that the larger number of patients in our study, differences in comorbidities, associated anomalies, and when interventions were offered account for the differences between these two studies. Nonetheless, based on our data and other published studies, it should also be clear that current neonatal intensive care can prolong the absolute length of survival for many infants with trisomy 13 or trisomy 18, and as a result, broadly applying the term “futility” to medical interventions for these infants is inappropriate.^9^

For these reasons, we find that it is incredibly important to look at outcome measures beyond absolute length of survival. Infants who survived beyond the first week of life had high rates of healthcare utilization throughout the remainder of their lifespan. Nearly half of infants discharged home were readmitted to the hospital at least one, with some children requiring numerous readmissions with a total time spent in the hospital lasting more than two years. Need for respiratory support, nutritional support, and invasive surgical procedures were relatively common. All of the children in our cohort that were discharged home received at least a portion of their nutrition through either temporary or surgically placed feeding tubes, and many children required home respiratory support, including tracheostomy and mechanical ventilation. In Japan, a national database was utilized to review hospital admissions and medical procedures in 133 patients with T13 and 438 patients with T18 over a three-year period. The majority of patients who were discharged required home medical care ranging from tube feedings to oxygen or mechanical ventilation.^36^ Other studies have highlighted that invasive surgical procedures are frequently being offered as treatment options for children with trisomy 13 or 18, with significant variability in patient outcomes and the impact of these procedures on overall survival.^6, 24, 27, 38, 40, 41^ Expected neurodevelopmental outcomes, level of functioning, and the degree of interaction that families have with their children are also critical factors that must be considered. Recent studies have begun to provide objective data on the long-term neurodevelopmental outcomes of children with T13 or T18, highlighting that these children do meet early developmental milestones and have meaningful interactions with their families.^42, 43^ Regardless of the treatment path chosen, the majority described the experiences with their child as positive and did not regret the decisions they made for either comfort measures or invasive interventions. ^8, 44^

The burden of congenital heart disease in infants and children with T13 and T18 is high; most surviving children with CHD confirmed by echocardiography ultimately developed symptoms requiring medical or surgical management. Deciding when surgical intervention is the best option for children with CHD and T13 or T18 remains a challenging decision for their families and pediatric care providers. A retrospective review for the Pediatric Care Consortium spanning 26 years reports that 58% of patients with T13 (mosaic or complete) and 57% of patients with T18 (mosaic or complete) underwent intervention for CHD. Of those, 27.6% (T13) and 13% (T18) died during that hospitalization and the median survival for those who survived to discharge was 14.8 years for patients with T13 and 16.2 years for patients with T18. There was variation in the cause of death, most being related to respiratory or cardiac issues.^45^ Some studies suggest that infants requiring earlier cardiac interventions have worse outcomes. Even with simple cardiac lesions, optimizing medical management to allow for complete repair when the infant is older may be beneficial.^46^ Another recent study suggests that cardiac surgery may be associated with decreased in-hospital mortality in a certain subset of patients. While congenital heart disease is a common finding in individuals with T13 or T18, the type of defect and approach to interventions, if offered, remains variable.^47^ A group recently attempted to address this issue by developing a prediction model for survival to 6 months. In their study, certain variables, including cardiac surgery, gastrostomy, parenteral nutrition, and mechanical ventilation, are predictive of survival to 6 months of age.^48^ This is consistent with data from our study. It appears that an individual approach is necessary in determining which patients could potentially benefit from invasive interventions, such as cardiac surgery. ^49^

Given the complexity of the decisions that need to be made by families and care providers, open and transparent discussions on the best available objective data are critical. With the changing approach to care for patients with T13 and T18, providers may feel distress or may personally disagree with the goals of care and treatment pathways families wish to pursue.^50^ It is important that providers understand the values and goals of the family, as well as the anticipated outcomes and disease trajectory, for each fetus or neonate with T13 or T18 to appropriately counsel families. In this cohort, less than half of families met with a pediatric provider or multidisciplinary fetal care team during their pregnancy. We hypothesize that the association between prenatal consultation with pediatric providers and lower termination rates (but lack of an association between this prenatal counseling and postnatal survival) reflects referral selection bias - families are making decisions about goals of care before ever meeting with pediatric care providers. Whether these decisions are being made based on counseling received from their primary obstetrician, consulting maternal fetal-medicine specialists, their own personal views, or perspectives obtained from their family and community support (e.g. family support groups, internet) remains unknown. Similarly, we speculate that our observation that prenatal consultation with a fetal cardiologist is associated with increased survival may also be the result of selection and referral bias – only families already highly motivated to pursue all interventions to maximize the length of survival of their children are being referred to and are meeting with a fetal cardiologist. However, as the majority of pregnancies resulted in a livebirth irrespective of the family’s ultimate goals of care, this may leave some families incompletely prepared for the experiences and decisions that they will face after birth. Only a slight majority of infants in this study had a birth plan in place at the time of delivery. Specialized centers now frequently have perinatal palliative care services available to women with a fetal life-limiting diagnoses, but many families are not referred to such programs.^4, 13^ Involvement of a palliative care team early on allows for the family to explore their values and views of quality of life. The unique and individualized approach to care allows for families and medical providers to work collaboratively to develop a care plan that aligns with the family’s wishes. ^12, 44, 51, 52 44, 51, 52^

Our study has several limitations related to the rarity of the two diagnoses and the retrospective nature of the study. While the integrated nature of newborn care across the Greater Cincinnati Region allows for a unique opportunity to perform outcomes research across a large population, and we made every possible effort to ascertain every prenatal and postnatal case of trisomy 13 or 18, we recognize that the data may not be complete. However, the number of patients identified for the study are consistent with what would be expected based on current literature for the number of pregnancies in the area, and the authors were able to find outcomes for all patients included in the study. Although retrospective cohort studies have their own limitations, due to the relatively rare nature of these diagnoses the time needed to complete a comparably sized prospective observational study would be impractical, and a controlled trial randomizing infants to different treatment pathways would be entirely unethical. Within the context of these limitations, this study provides key insights into the prenatal, perinatal and postnatal outcomes of fetuses and infants with trisomy 13 and 18, and should serve as a useful frame-of-reference to facilitate counseling the families of children with these diagnoses. Future research is needed to better understand what information families are utilizing to establish goals of care for their children, particularly during the prenatal period, to better understand the long-term medical and developmental outcomes for children after the first month of life, and the relationship between these children and their families.

## Data Availability

All data produced in the present study are available upon reasonable request to the authors

## List of Abbreviations

CHD: Congenital Heart Disease
ELBW: Extremely Low Birthweight
ELGAN: Extremely Low Gestational Age Neonate
NICU: Neonatal Intensive Care Unit
T13: Trisomy 13
T18: Trisomy 18
VLGAN: Very Low Gestational Age Neonate

## Acknowledgements

We would like to thank Dr. Jeffrey Whitsett and Dr. James Greenberg for their critical review of this manuscript. D.T.S. was supported by NIH R01HL156860 during the study period.

## Supplemental Data

**Supplemental Table 1.**
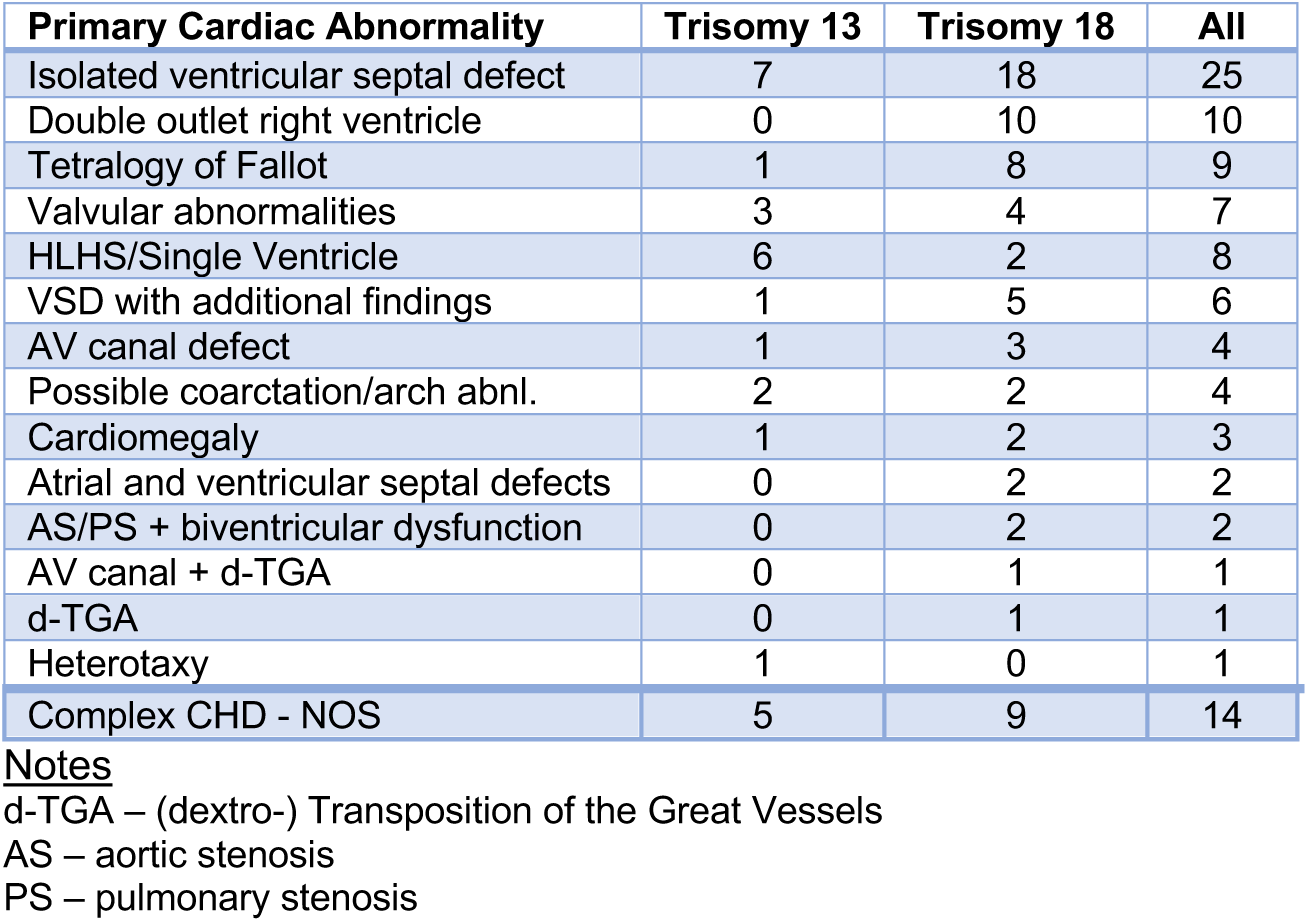
Prenatal Cardiac Findings.

| Primary Cardiac Abnormality | Trisomy 13 | Trisomy 18 | All |
| --- | --- | --- | --- |
| Isolated ventricular septal defect | 7 | 18 | 25 |
| Double outlet right ventricle | 0 | 10 | 10 |
| Tetralogy of Fallot | 1 | 8 | 9 |
| Valvular abnormalities | 3 | 4 | 7 |
| HLHS/Single Ventricle | 6 | 2 | 8 |
| VSD with additional findings | 1 | 5 | 6 |
| AV canal defect | 1 | 3 | 4 |
| Possible coarctation/arch abnl. | 2 | 2 | 4 |
| Cardiomegaly | 1 | 2 | 3 |
| Atrial and ventricular septal defects | 0 | 2 | 2 |
| AS/PS + biventricular dysfunction | 0 | 2 | 2 |
| AV canal + d-TGA | 0 | 1 | 1 |
| d-TGA | 0 | 1 | 1 |
| Heterotaxy | 1 | 0 | 1 |
| Complex CHD - NOS | 5 | 9 | 14 |

**Supplemental Table 2.** Factors Associated with Spontaneous Loss.

|  | Spontaneous Loss<br>(n = 32) | Liveborn<br>(n = 72) | P-Value |
| --- | --- | --- | --- |
| **Maternal Age** (median) | 34 | 33 | NS |
| **Gravida** (median) | 2 | 3 | NS |
| **Fetal Gender** (Male) | 59% | 36% | < 0.05 |
| **Trisomy Type** (T13) | 25% | 35% | NS |
| **Organ Systems Affected\*** | 3 | 3 | NS |
| **Hydrops\*** | 16% | 3% | < 0.05 |
| **CHD\*** | 61% | 88% | < 0.01 |

**Supplemental Table 3.** Association Between Antenatal Counseling & Pregnancy Outcome.

|  | Liveborn/Spontaneous Loss<br>(n = 104) | Termination<br>(n = 21) | P-Value |
| --- | --- | --- | --- |
| **Neonatology Consult** | 46 (48%) | 1 (4.8%) | < 0.001 |
| **Fetal Care Consult** | 18 (19%) | 2 (10%) | NS |
| **Cardiology Consult** | 37 (39%) | 1 (4.8%) | < 0.01 |
| **Palliative Care Consult** | 56 (58%) | 1 (4.8%) | < 0.001 |

**Supplemental Table 4.**
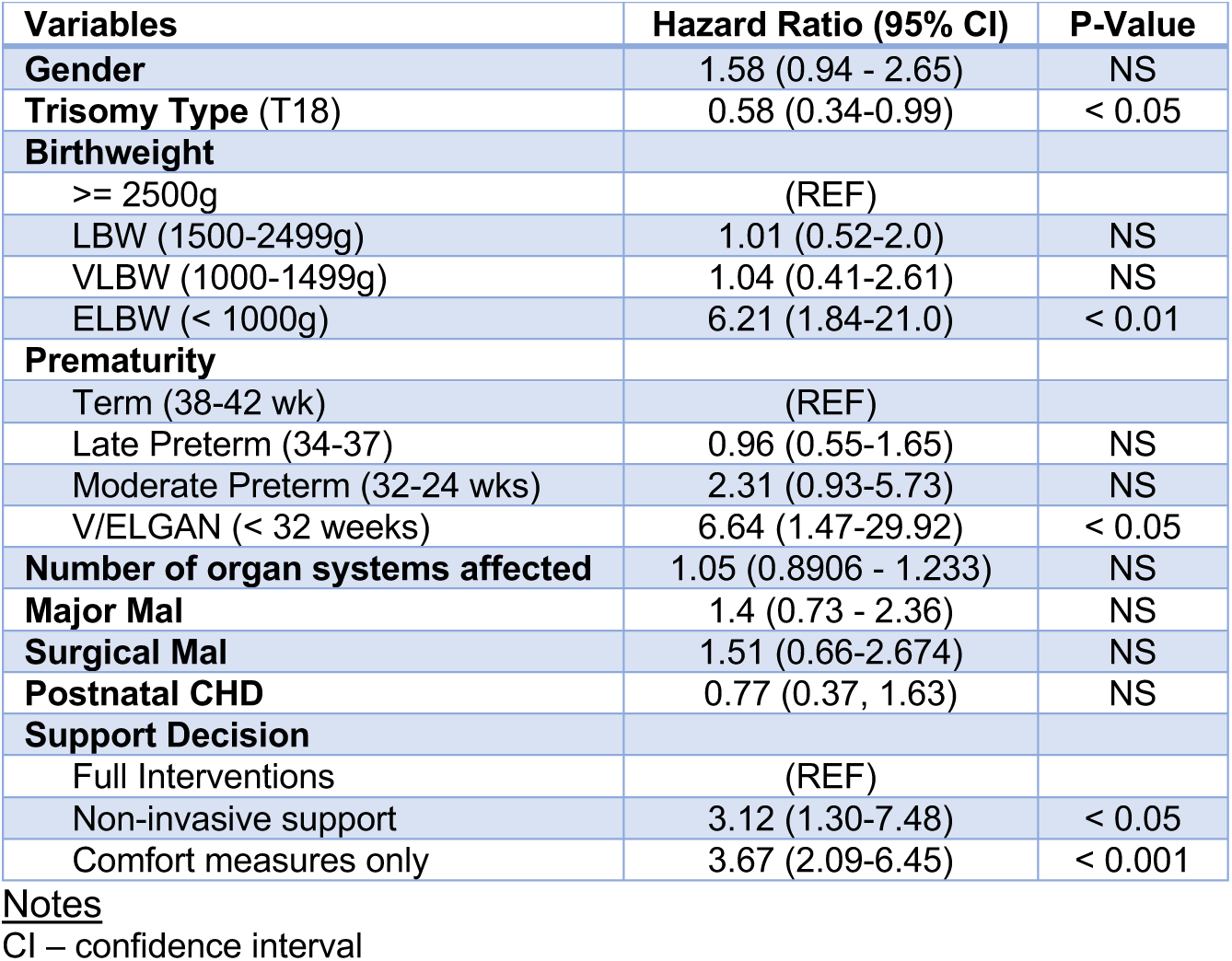
Factors Associated with Survival (Univariate Analysis)

**Supplemental Table 5.**
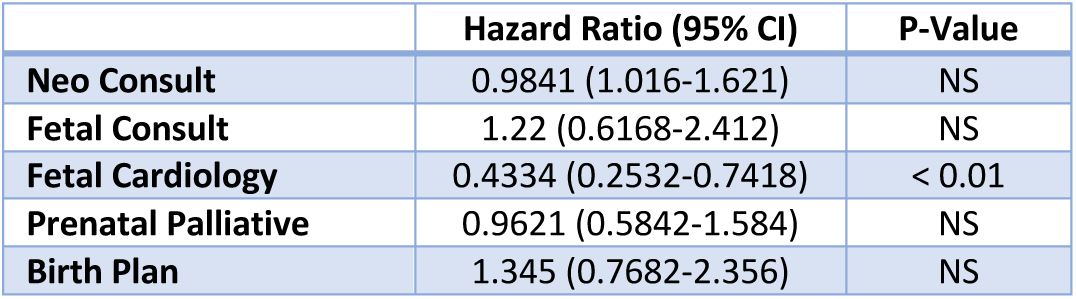
Prenatal Consultation & Postnatal Survival.

**Supplemental Table 6.** Medical Interventions, Hospitalizations & Survival of Liveborn Infants.

|  | **Trisomy 13**<br>(n = 25) | **Trisomy 18**<br>(n = 47) | **All**<br>(n = 72) |
| --- | --- | --- | --- |
| **Admitted to the NICU** | 9 (36%) | 23 (49%) | 32 (44%) |
| **Discharged home** | 8 (32%) | 24 (51%) | 32 (44%) |
| Readmitted to the Hospital* | 3 (38%) | 12 (50%) | 15 (47%) |
| **Respiratory** |  |  |  |
| Apnea | 20 (80%) | 43 (91%) | 63 (88%) |
| Respiratory Support | 13 (52%) | 21 (45%) | 34 (47%) |
| Mechanical Ventilation | 9 (36%) | 8 (17%) | 17 (24%) |
| **Congenital heart disease (any)** | 19 (76%) | 42 (89%) | 61 (85%) |
| **Cardiology** |  |  |  |
| Post-natal echo | 10 (40%) | 25 (53%) | 35 (49%) |
| Symptomatic Needing Meds* | 6 (60%) | 18 (72%) | 24 (69%) |
| Pulmonary Hypertension* | 5 (50%) | 13 (52%) | 18 (51%) |
| **Surgical Interventions** | 3 (12%) | 17 (36%) | 20 (28%) |
| **Approach to Care** |  |  |  |
| Comfort Care Only | 12 (48%) | 22 (47%) | 34 (47%) |
| Non-invasive Interventions | 11 (44%) | 20 (43%) | 31 (43%) |
| Invasive Interventions | 2 (8%) | 5 (11%) | 7 (10%) |
| **Do Not Resuscitate Orders** | 24 (96%) | 37 (79%) | 61 (85%) |
| At Birth* | 14 (58%) | 23 (62%) | 37 (61%) |
| After a Trial of Interventions* | 7 (29%) | 8 (22%) | 15 (29%) |
| Just Before Death* | 3 (13%) | 6 (16%) | 9 (15%) |
| **Died** | 23 (92%) | 40 (85%) | 63 (88%) |
| At Home* | 5 (22%) | 12 (30%) | 17 (27%) |
| In the Hospital* | 18 (78%) | 27 (68%) | 45 (71%) |
| In the Ambulance* | 0 | 1 (2%) | 1 (2%) |
| **Alive** | 2 (8%) | 7 (15%) | 9 (13%) |
| Median (range) | 584 (531-637) | 1394 (318-2762) | 1366 (318-2762) |

Notes
\* Note that the relevant denominator was used to calculate percentages for each category, which may not equal all liveborn infants with T13 or T18 (e.g. percentage of infants readmitted to the hospital is referenced to the total number of infants discharged home)

**Supplemental Table 7.** Surgical Procedures for Children with Trisomy 13 or 18.

| Organ System & Surgery Type | T13 | T13 | T13 | T18 | T18 | T18 |
| --- | --- | --- | --- | --- | --- | --- |
|  | # Surgeries | # Infants | Age Range | # Surgeries | # Infants | Age Range |
| **Cardiology** |  |  |  |  |  |  |
| Major | 0 | 0 | n/a | 5 | 5 | 49-283 |
| Minor | 0 | 0 | n/a | 2 | 2 | 79-228 |
| **Gastrointestinal** |  |  |  |  |  |  |
| Major | 3 | 2 | 23-246 | 21 | 11 | 1-1526 |
| Minor | 6 | 1 | 72-325 | 23 | 12 | 28-1526 |
| **Ears/Nose/Throat** |  |  |  |  |  |  |
| Major | 2 | 2 | 174-246 | 8 | 8 | 28-1586 |
| Minor | 1 | 1 | 627 | 6 | 5 | 521-1304 |
| **Plastics** |  |  |  |  |  |  |
| Major | 1 | 1 | 261 | 2 | 2 | 655-1613 |
| **Neurosurgery** |  |  |  |  |  |  |
| Major | 0 | 0 | n/a | 3 | 1 | 27-50 |
| **Other** |  |  |  |  |  |  |
| Major | 0 | 0 | n/a | 8 | 3 | 134-2173 |
| Minor | 0 | 0 | n/a | 13 | 7 | 5-1526 |
| **Specific Surgeries By Trisomy Type** |  |  |  |  |  |  |
| **Surgery** | **T13** | **T18** | **Surgery** | **T13** | **T18** |  |
| **Gastrointestinal** |  |  | **Ears/Nose/Throat** |  |  |  |
| G-tube | 3 | 15 | Tracheostomy | 2 | 8 |  |
| GJ/GJ revision | 2 | 6 | Myringotomy tubes | 1 | 4 |  |
| Nissen | 0 | 4 | Adenoidectomy/ Tonsillectomy | 0 | 2 |  |
| Ladd's | 1 | 3 | **Plastics** |  |  |  |
| Hepatic resection/cholecystectomy | 0 | 1 | Cleft lip/palate | 1 | 1 |  |
| Bowel Resection | 2 | 3 | Craniotomy | 0 | 1 |  |
| Omphalocele closure | 0 | 1 | **Neurosurgery** |  |  |  |
| Meckel's | 0 | 1 | EVD/VP shunt | 0 | 2 |  |
| Umbilical hernia | 1 | 1 | MMC | 0 | 1 |  |
| TE fistula | 0 | 2 | **Other** |  |  |  |
| Pyloroplasty | 0 | 1 | Central lines | 0 | 3 |  |
| Paraesophageal hernia | 0 | 3 | Aortopexy | 0 | 1 |  |
| Embolize liver tumor/liver biopsy | 0 | 3 | Diaphragm plication | 0 | 1 |  |
| **Cardiology** |  |  | ECMO (1 failed cannulation) | 0 | 2 |  |
| Cath (1 attempted VSD/ASD closure) | 0 | 3 | Botox | 0 | 5 |  |
| ASD/VDS/PDA closure | 0 | 4 | Chest exploration | 0 | 2 |  |

**Supplemental Table 8.** Postnatal Echocardiogram Findings.

| Cardiac Lesion | Trisomy 13<br>(n = 10) | Trisomy 18<br>(n = 25) | Totals<br>(n = 35) |
| --- | --- | --- | --- |
| Ventricular Septal Defect | 7 | 23 | 30 |
| Atrial Septal Defect/Patent Foramen Ovale | 5 | 16 | 21 |
| Valvular abnormalities | 4 | 14 | 18 |
| Patent Ductus Arteriosus | 5 | 7 | 12 |
| Double Outlet Right Ventricle | 0 | 4 | 4 |
| Tetralogy of Fallot | 3 | 1 | 4 |
| Coarctation/Arch abnormalities | 1 | 2 | 3 |
| d-Transposition of the Great Vessels | 0 | 2 | 2 |
| Hypoplastic Left Heart Syndrome | 1 | 0 | 1 |
| Truncus arteriosus | 0 | 0 | 0 |
| AV Canal Defect | 0 | 0 | 0 |

**Supplemental Figure 1.**
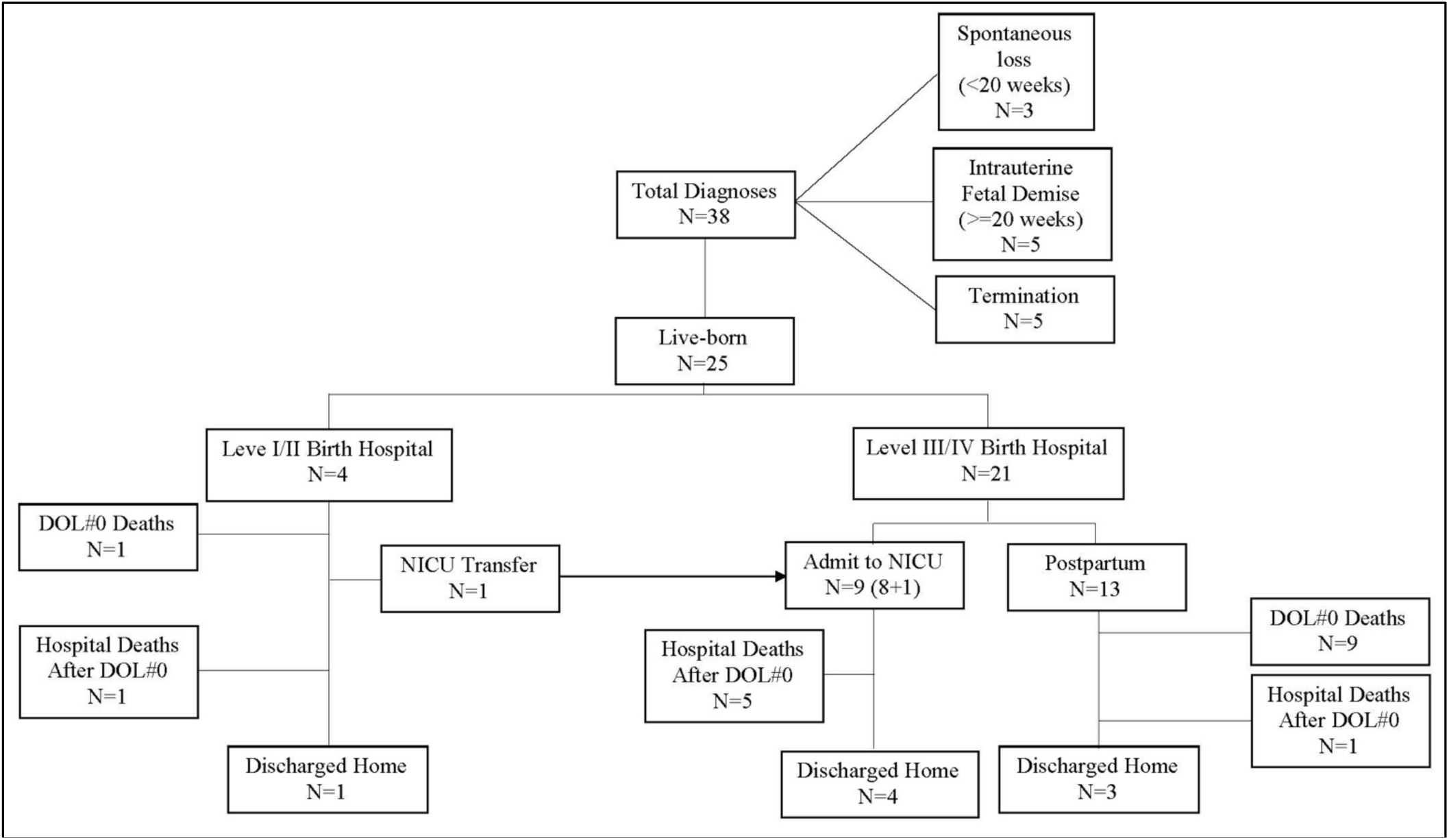
Postnatal Outcomes Flow Diagram for Trisomy 13. The number of pregnancies and liveborn infants with trisomy 13 enrolled in the study are outlined.

**Supplemental Figure 2.**
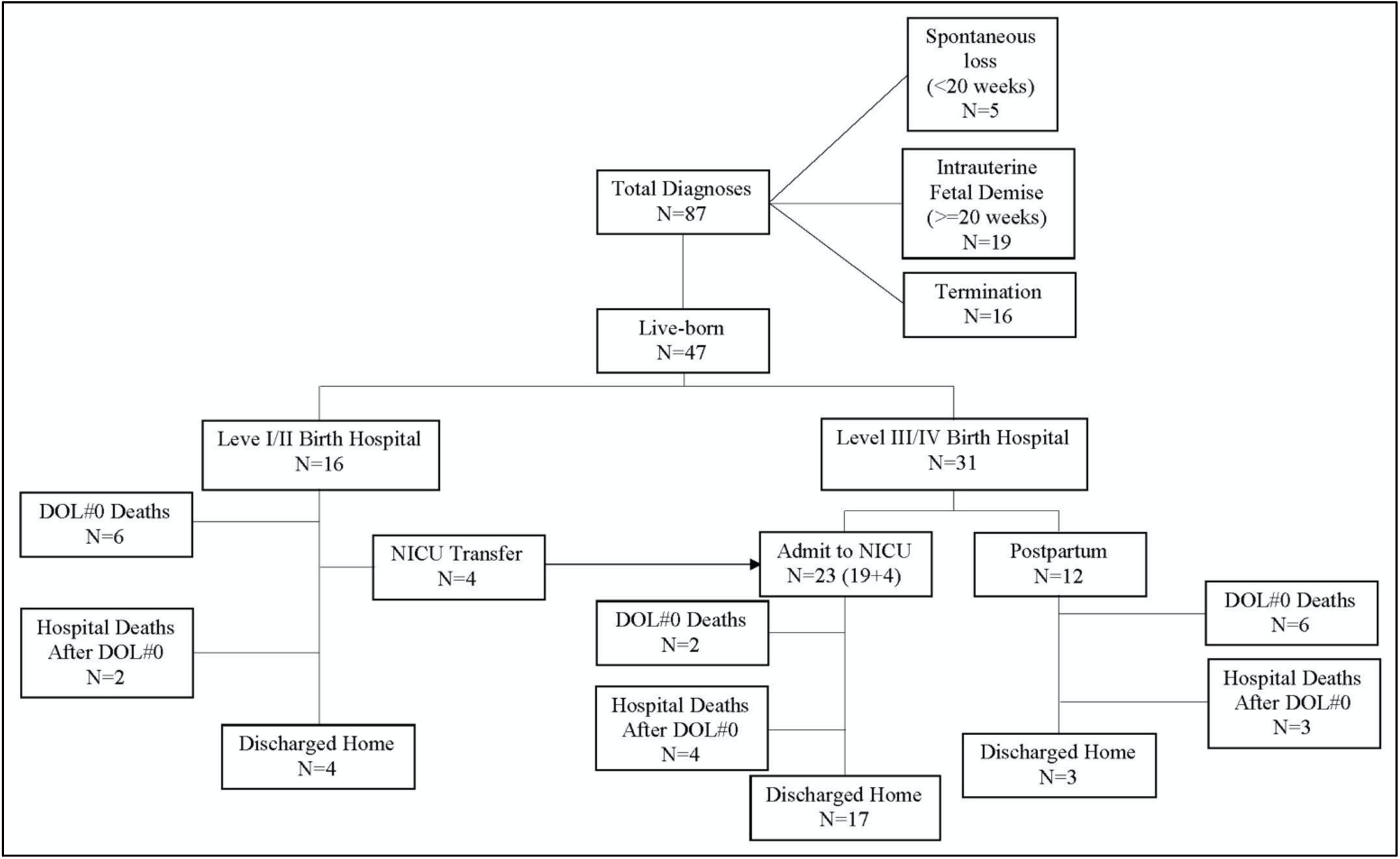
Postnatal Outcomes Flow Diagram for Trisomy 18. The number of pregnancies and liveborn infants with trisomy 18 enrolled in the study are outlined.

**Supplemental Figure 3.**
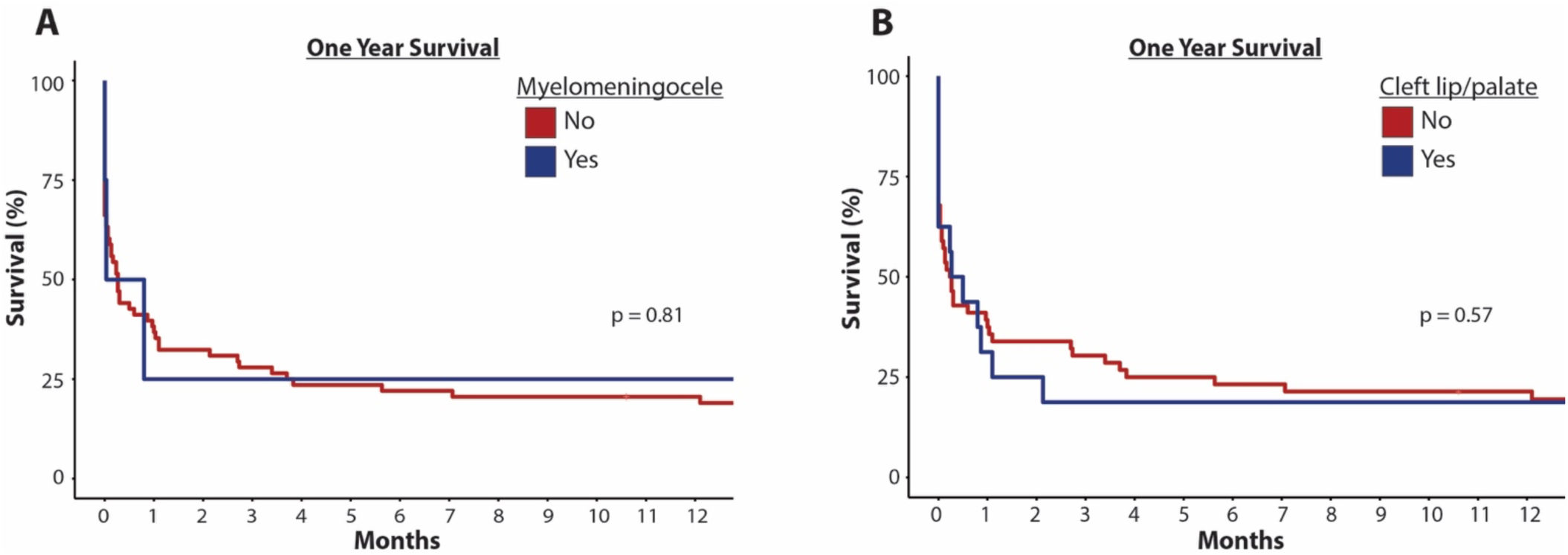
Survival for Children with Trisomy 13 or 18 with myelomeningocele or cleft lip/palate. (**A-B**) Kaplan-Meier survival plots are shown for children with T13 or T18 with myelomeningocele (A) or cleft lip/palate (B).

